# Assessing the Impact of Framework Agreements on the Availability of Essential Medicines: A Case Study of 65 Essential Medicines at the Upper East Regional Medical Stores, Ghana

**DOI:** 10.1101/2025.09.10.25335483

**Authors:** Abdalla Salia, Baba Seidu Issifu, Kwaku Gyamfi Oppong

## Abstract

**Background:** Access to Essential Medicines stands as a vital foundation to public health service delivery in any well-performing health system. In 2017, Ghana’s Ministry of Health established the Framework Agreement system which selects prequalified suppliers to deliver 54 Essential Medicines (now 65 Essential Medicines) to Regional Medical Stores and Teaching Hospitals to address supply chain inefficiencies and maintain continuous availability.

**Objectives:** The study assessed the impact of these Framework Agreements on medicine availability at the Upper East Regional Medical Stores (UERMS), focusing on effectiveness, implementation challenges and stakeholder perceptions.

**Methods:** A mixed-methods cross-sectional study design was employed. Quantitative data were collected through retrospective analysis of stock records from 2015 to 2024, comparing availability and lead times before and after implementation, while qualitative data were gathered through semi-structured interviews with key stakeholders.

**Results:** Quantitative data showed that prior to implementation, stock availability of these 65 Essential Medicines was 100%, with average lead times of 21–22 days. However, after implementation (2017–2024), medicine availability declined, and lead times increased by more than 40%, undermining the system’s objective of efficiency. Inconsistencies in supply were observed. Qualitative data showed that supplier performance faced system-level obstacles, including restricted emergency procurement flexibility, communication breakdowns between procurement entities and suppliers, and delayed payments, leading suppliers to lose commitment.

**Conclusion:** Framework Agreements demonstrate potential for better procurement transparency and cost-effectiveness yet its existing implementation model negatively impacts medicine availability in the Upper East Region. The study recommends revision to allow emergency procurement, strengthen supplier performance monitoring, decentralise some procurement responsibilities, improve communication and ensure timely payments.

## INTRODUCTION

Access to Essential Medicines is a major factor influencing health outcomes and a fundamental component of healthcare delivery.^17^ The WHO defines Essential Medicines as those satisfying the priority healthcare needs of a population, selected based on disease frequency, efficacy, safety, and cost-effectiveness.^16^ These medicines form the backbone of healthcare, particularly in public health facilities, making their supply a top priority for the Ministry of Health (MOH) and Ghana Health Service (GHS).^5,6,14^ Access to medicines is a fundamental human right and a key building block of effective health systems.^10,11^ However, records indicate that supplies in Ghana’s public health institutions are often below the WHO-recommended threshold of 80%, reflecting challenges in meeting patient needs.^6^

Framework Agreements (FWAs) are long-term memoranda defining the terms for recurring smaller purchasing orders.^1^ They aim to increase medicine availability, simplify procurement, and ensure cost-effectiveness.^15^ Yet, their effectiveness at ensuring continuous supply at regional and institutional levels, particularly in the Upper East Regional Medical Stores (UERMS), remains uncertain, with stakeholders questioning whether FWAs truly improve pharmaceutical supply.

The FWA system relies on pre-negotiated contracts with selected suppliers to provide essential medicines over a defined period, mitigating procurement inefficiencies, ensuring quality, and stabilizing prices.^4^ After the 2015 Central Medical Stores fire, Ghana developed a Supply Chain Master Plan implemented through collaboration between MOH and the Global Fund, including Framework Contracting/Agreement (FWA), Last Mile Distribution (LMD), GhiLMIS, and Warehouse Optimisation. The FWA was designed to address stockouts, high acquisition costs, and supply chain inefficiencies.^8^ Under the system, selected suppliers provide 65 Essential Medicines— including antibiotics, antihypertensives, antidiabetics, haematinics, antiulcers, antihistamines, vaccines, immunoglobulins, ophthalmics, and other essential drugs.^3^ Nevertheless, studies indicate potential drawbacks, such as delayed supplier response, reduced procurement flexibility, and sporadic stockouts.^3^

In the Upper East Region, geographical and infrastructural limitations make access to healthcare challenging, making consistent Essential Medicines supply critical.^12^ Irregular supply may increase morbidity and mortality, particularly among vulnerable populations dependent on public health facilities.^17^ While centralised procurement through FWAs can be efficient, reliance on pre-selected suppliers can extend stockouts during manufacturing delays or logistical challenges.^2^ Understanding these issues is essential for planning improvements in procurement and ensuring continuous medicine availability.

Achieving universal health coverage and improving patient outcomes depends on uninterrupted Essential Medicines supply.^17^ Preliminary data suggest that UERMS faces challenges in maintaining sufficient stocks of the 65 Essential Medicines, affecting local healthcare delivery.^2^ FWAs, while providing long-term contracts, do not fully account for demand fluctuations from disease outbreaks or seasonal consumption variations.^7^ Moreover, regional medical stores’ limited ability to engage in contingency procurement outside FWAs may exacerbate supply disruptions.^12^ Therefore, evaluating the efficiency of the FWA procurement method and its impact on healthcare delivery in the Upper East Region is critical.

## METHODS

### Study Design

This study employed a descriptive cross-sectional design using both quantitative and qualitative methods (mixed methods approach). The quantitative component involves a retrospective review of stock records to assess the availability of the 65 Essential Medicines, while the qualitative component explores perceptions and experiences of key stakeholders regarding the implementation and effectiveness of Framework Agreements.

### Study Area

The study was conducted in the Upper East Region of Ghana, focusing on the Upper East Regional Medical Stores (UERMS) located in Bolgatanga, and selected public health facilities (District Hospitals and the Regional Hospital) that are directly supplied by the Upper East Regional Medical Stores. The region is characterised by a mix of urban and rural populations and experiences unique challenges in health logistics due to its geographic location and infrastructural constraints.

### Study Site

The study was conducted in the Upper East Regional Medical Stores, a unit of the Upper Regional Health Directorate located in Bolgatanga. The Upper East Regional Medical Stores (UERMS) operates as a vital supply chain element for health services in Upper East Region and it is under the Upper East Regional Health Directorate’s governance. The UERMS serves as the essential hub for health commodity storage, distribution, and management throughout the Upper East Region. The UERMS functions as a fundamental logistical hub which ensures and maintains proper pharmaceutical product, non-medicine consumables, and medical equipment availability for all public health facilities and some private health facilities in the region. The core mandate of the Upper East Regional Medical Stores includes the receipt, safe storage, and timely distribution of medical commodities to health facilities. The facility manages essential medicines and vaccines alongside laboratory reagents and medical devices together with non-medicine consumables. The UERMS currently serves 15 District and Municipal Health Directorates, 1 Regional Hospital, 8 District Hospitals, 14 CHAG Health facilities, 12 Private Hospitals, 76 Health Centres, and 240 CHPS. It performs its operations based on national health policy directives and Ministry of Health’s guidance and it maintains close partnerships with the Central Medical Stores as well as Non-Governmental Organisations and accredited private sector suppliers. The UERMS maintains operational efficiency through its implementation of diverse supply chain management and logistics practices. The facility maintains inventory control systems through the Ghana Integrated Logistics Management Information System (GhiLMIS) and conducts regular stock audits while preserving temperature-sensitive products through cold chain systems and follows Good Storage Practices (GSP) standards. The facility operates with separate units dedicated to warehousing, storage, inventory control, quality assurance, and dispatch operations. The UERMS again supports vertical public health programs such as Expanded Program on Immunisation (EPI), Tuberculosis, Malaria, HIV/AIDS and Family Planning through commodity management and distribution services. Through its activities, the UERMS supports the Ghana Health Service’s objectives to enhance healthcare delivery while promoting universal health coverage and equity in the region. The Upper East Regional Medical Stores maintains its strategic position as the central healthcare delivery point to evaluate the effects of Framework Agreements on essential medicine availability.

### Study population

The target population consists of three main groups: Supply Chain Officers at the UERMS who manage and supervise the receipt, storage, and distribution of Essential Medicines, Pharmacists at the Regional and District Hospitals within the Upper East Region who receive supplies from the UERMS, and Representatives of selected pharmaceutical suppliers who have been awarded Framework Agreements to supply the 65 Essential Medicines.

### Inclusion and Exclusion Criteria

Inclusion criteria include Pharmacists with at least one year of experience at their current facility, Supply Chain Practitioners who are directly involved in procurement, ordering, or inventory management, Pharmaceutical Suppliers who have active contracts under the Framework Agreement system and Pharmaceutical Suppliers who have stopped supplying the UERMS under the Framework Agreement system. Exclusion criteria include interns or temporary staff without adequate knowledge of procurement processes, and Pharmaceutical Suppliers not included in the Framework Agreement for the selected medicines.

### Sample Size and Sampling Techniques

A purposive sampling technique was used to select participants based on their relevance to the study. The proposed sample size includes 1 Regional Medical Store (UERMS) staff, 8 District Hospital Pharmacists, 1 Regional Hospital Pharmacist and 10 Representatives of Pharmaceutical Suppliers with active Framework Agreements as well as Suppliers who have stopped supplying the UERMS under the Framework Agreement. This yields an estimated sample size of 20 participants, which is sufficient for achieving data saturation in qualitative studies and allows meaningful quantitative summary of availability levels.

### Data Collection Methods

#### Quantitative Data

A retrospective review of stock records was conducted from 01/04/2025 to 30/04/2025, to assess the availability of the 65 Essential Medicines over a 10-year period (2015–2024), covering 2 years before and 8 years after the implementation of the Framework Agreement. Data was extracted using a structured template and analysed to identify trends and patterns in medicine availability before and after the framework agreement policy implementation.

#### Qualitative Data

Semi-structured interviews were conducted with the selected pharmacists, UERMS staff, and supplier representatives from 01/05/2025 to 31/05/2025. The interview guide explored perceptions of framework agreement implementation, perceived impact on availability and delivery timelines, challenges experienced and recommendations for improvement.

### Data Analysis

#### Quantitative Data Analysis

Quantitative data was entered into Microsoft Excel and analysed using SPSS. Descriptive statistics were generated to show availability trends.

#### Qualitative Data Analysis

Thematic content analysis was used to examine open-ended responses. Responses were transcribed and then initial codes assigned to meaningful phrases and statements. Then these codes were grouped into broader themes corresponding to the study objectives including availability of Essential Medicines, Framework Agreement challenges, supply delays and procurement flexibility. This approach allows for a comprehensive understanding of the experiences of stakeholders about the Framework Agreement’s influence on the availability of the 65 Essential Medicines at the hospitals and Upper East Regional Medical Stores.

#### Ethical Considerations

Ethical clearance was obtained from the Committee on Human Research Publication and Ethics (CHRPE), KNUST, with reference number CHRPE/AP/437/25. Participation in the study was voluntary. All participants provided written informed consent before data collection. Data confidentiality and anonymity was strictly maintained.

## RESULTS

### Quantitative Findings

From 2015–2016 (pre-Framework Agreement), stock availability for the 65 essential medicines at the Upper East Regional Medical Stores (UERMS) was consistently 100%, with mean lead times of 21–22 days. Post-implementation (2017–2024), availability dropped, with frequent stock-outs, and mean lead times increased by more than 40% as shown in Table 1.

**Table 1.** Stock availability and lead times before and after Framework Agreement implementation.

| YEAR | PERCENTAGE STOCK AVAILABILITY (%) | AVERAGE LEAD TIME (DAYS) | FRAMEWORK AGREEMENT STATUS |
| --- | --- | --- | --- |
| 2015 | 100 | 20 | INACTIVE |
| 2016 | 100 | 21 | INACTIVE |
| 2017 | 82 | 23 | ACTIVE |
| 2018 | 71 | 24 | ACTIVE |
| 2019 | 55 | 26 | ACTIVE |
| 2020 | 75 | 27 | ACTIVE |
| 2021 | 83 | 28 | ACTIVE |
| 2022 | 58 | 29 | ACTIVE |
| 2023 | 69 | 28 | ACTIVE |
| 2024 | 52 | 29 | ACTIVE |

Year-by-year data indicated that the steepest decline in availability occurred in 2017– 2019, and Lead times showed the greatest increase during 2021 to 2024 shown in **Fig 1**.

**Fig 1.**
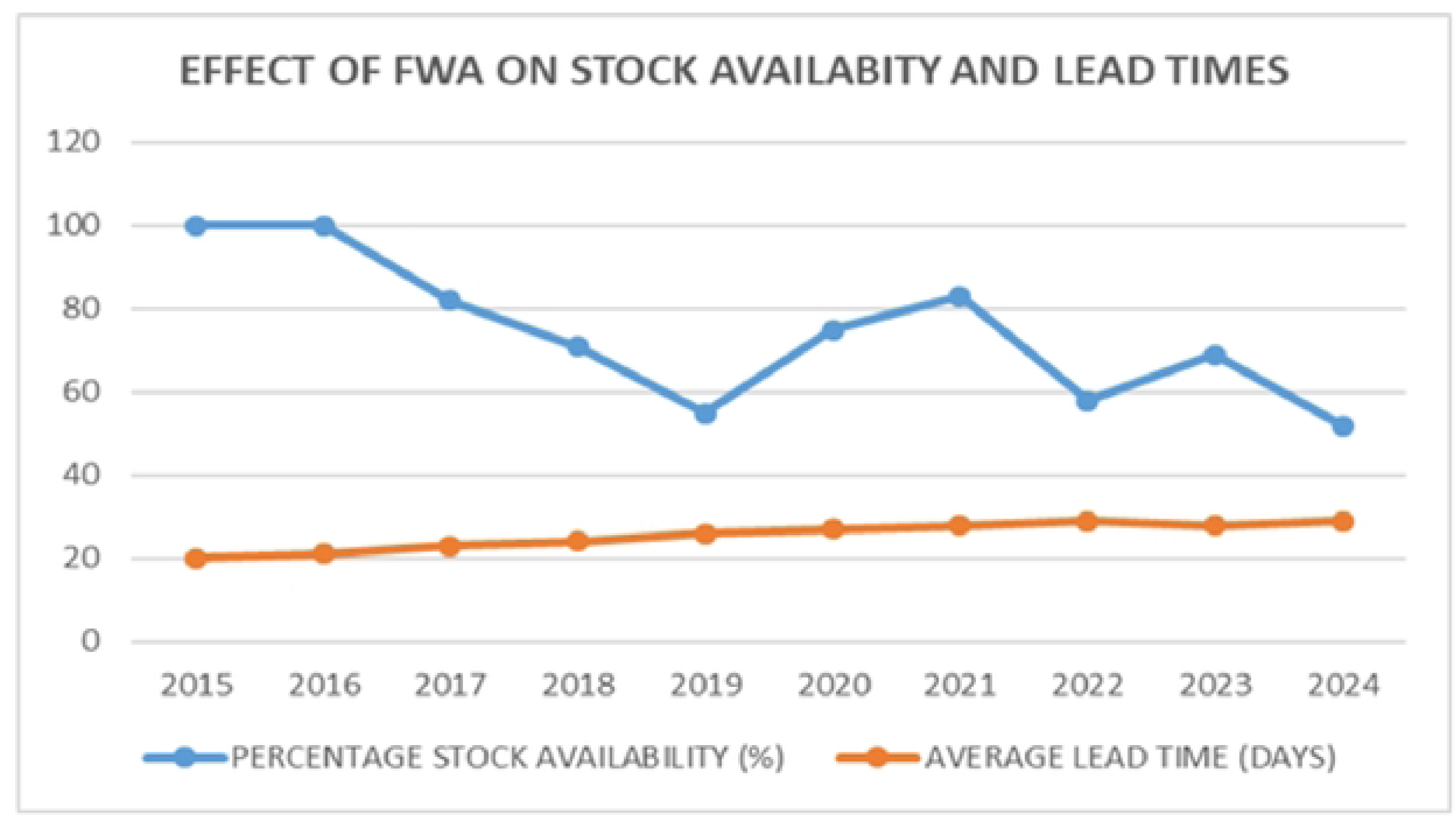
Effect of Framework Agreements on Stock Availability and Lead Times

### Qualitative Findings

**Table 2.**
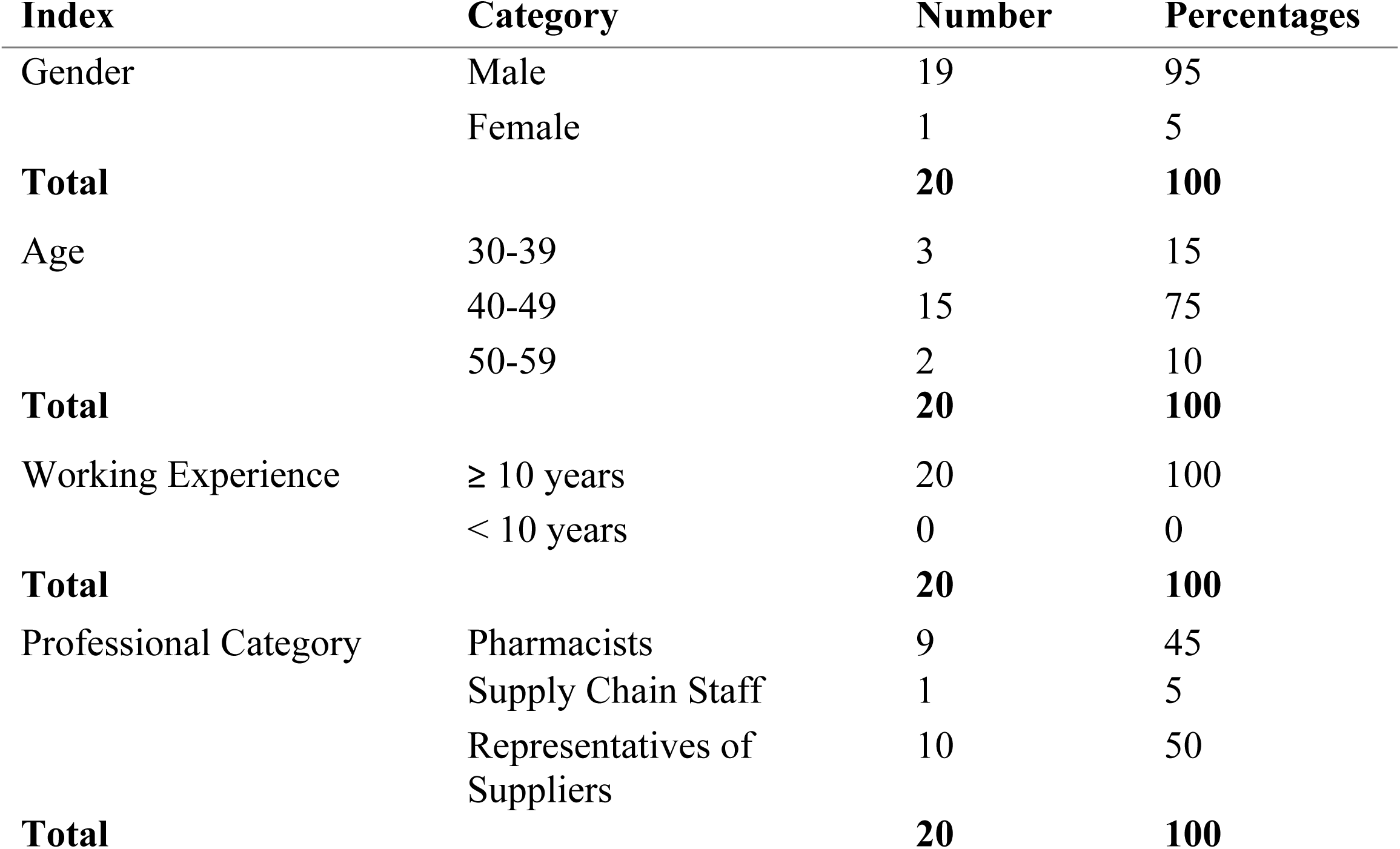
Demographic profile of respondents.

| Index | Category | Number | Percentages |
| --- | --- | --- | --- |
| Gender | Male | 19 | 95 |
|  | Female | 1 | 5 |
| **Total** |  | **20** | **100** |
| Age | 30-39 | 3 | 15 |
|  | 40-49 | 15 | 75 |
|  | 50-59 | 2 | 10 |
| **Total** |  | **20** | **100** |
| Working Experience | ≥ 10 years | 20 | 100 |
|  | < 10 years | 0 | 0 |
| **Total** |  | **20** | **100** |
| Professional Category | Pharmacists | 9 | 45 |
|  | Supply Chain Staff | 1 | 5 |
|  | Representatives of Suppliers | 10 | 50 |
| **Total** |  | **20** | **100** |

### Stakeholders Take on the Framework Agreement Policy

#### Policy Content

Out of the 20 respondents from the 3 main groups of population, 2 respondents constituting 10% were aware of the policy but had no idea about its content whereas 18 respondents constituting 90% were both aware of and have knowledge of the content of the policy as indicated in **Table 3**. Though some were aware of the policy, they were unable to articulate its parts, goals and objectives.

**Table 3.** Respondents awareness of the Framework Agreement Policy.

| Indicators | Number | Percentage |
| --- | --- | --- |
| Not aware of the policy | 0 | 0 |
| Aware of the policy but have no knowledge of its content | 2 | 10 |
| Aware of the policy and have knowledge of its content | 18 | 90 |
| **Total** | **20** | **100** |

#### Policy Content

According to the study’s findings, a number of issues made Framework Agreement Policy’s rollout and execution necessary. Among these are reduction in administrative bottlenecks and enhancing equity, effectiveness and quality of care. Other technical issues included reducing procurement corruption, cutting down the prices of the medicines procured under this policy, addressing the issues of low staff capacity, particularly at the regional level and a lack of funding at the lower levels to engage in large volumes of procurement to enjoy economies of scale. The research on policy analysis supports these contextual factors by identifying sector reforms, administrative and technical bottlenecks, economic factors, traditional and sociocultural factors, environmental factors and demographic shifts as key policy determinants.^9^

#### Policy Implementation and Challenges

The implementation of the Framework Agreement (FWA) policy in the Upper East Region has encountered notable challenges. While the policy seeks to improve efficiency and transparency in public sector medicine procurement, its rollout has been marked by inconsistencies. A Supply Chain Manager at the Regional Medical Stores reported delays in supplier deliveries and difficulty in navigating the new centralised system, leading to inconsistent availability of Essential Medicines. A principal pharmacist at a district hospital stated, *“The idea is good, but the way it is being rolled out makes it difficult for us to maintain consistent availability of essential medicines.”*

Suppliers under the framework agreement also highlighted bottlenecks, citing poor coordination and a lack of engagement with the Ghana Health Service after contracts were awarded. Some expressed concerns about delayed payments for medicines already supplied. A supplier representative noted, *“We stopped supplying the various Regional Medical Stores across the country under the Framework Agreement because of delays in payment.”*

A Supply Manager explained that inadequate financial resources make it difficult to pay suppliers on time, largely due to erratic and delayed NHIS reimbursements, which in turn affects facilities’ ability to settle debts. The rigid nature of the policy also limits flexibility during emergencies or stockouts. A Senior Pharmacist asserted, *“In cases where the contracted suppliers fail to supply the Upper East Regional Medical Stores which has been our primary source of procurement, we are not allowed to procure from other sources even in urgent situations because the Regional Medical Stores would not issue certificate of non-availability to us.”*

Another issue raised by the Supply Manager was the absence of robust monitoring and evaluation systems. Without a standardised reporting mechanism for delays or defaulting suppliers, facilities were often forced to wait longer or resort to informal channels, undermining the purpose of the framework agreement.

## DISCUSSIONS, CONCLUSION AND RECOMMENDATIONS

### Discussions

From the quantitative findings, the implementation of the Framework Agreement (FWA) policy in 2017 led to a substantial reduction in stock availability. Stock availability reached 100% from the year 2015 to 2016 before the policy took effect. The Framework Agreement activation in 2017 resulted in stock availability decreasing to 82% and subsequently to 52% by 2024. The Framework Agreement’s current structure demonstrates an unsuccessful achievement of its purpose to sustain or enhance essential medicine supply levels. The supply chain efficiency decreased when lead times rose from 20–21 days in the 2015–2016 period (Pre-FWA period) to 29 days in 2024. The main reason for adopting FWAs was to enhance efficiency and decrease procurement delays yet the data shows the opposite effect. The data shows that the FWA implementation created new procurement inefficiencies because of bureaucratic rigidity, supplier non-compliance and centralization challenges. The policy aimed to create standard procurement procedures while shortening delivery times through prequalified pharmaceutical suppliers. The opposite outcome has occurred instead of the expected results. The lead times have grown longer throughout the years until they reached their highest point at 29 days during the years 2022 and 2024. The Framework Agreement has made stock availability more unpredictable which directly affects medicine security and public health results. The pre-FWA years maintained 100% stock availability but the FWA years showed a significant decline which suggests suppliers cannot meet demand or the system lacks proper enforcement for timely delivery. The study therefore shows that Framework Agreements have significant impact on the availability of the 65 Essential Medicines at the Upper East Regional Medical Stores and this impact, specifically, appears to be negative.

From the qualitative findings, the study discovered multiple obstacles which impede the Framework Agreement implementation in the Upper East Region. The main obstacle is the lack of coordination between the Ghana Health Service and contracted suppliers. Suppliers identified inadequate engagement together with delayed payments and restricted Ghana Health Service responsiveness as essential barriers to their work. This aligns with procurement literature, which emphasizes that successful supplier partnerships require ongoing communication, timely payments and mutual accountability.^13^ A supplier representative explained that they left the Framework Agreement because of ongoing payment delays. Health facility managers expressed their agreement with the problem of recurrent medicine stockouts and non-issuance of Certificates of Non-Availability which would have allowed them to purchase medicines outside the agreement. The Framework Agreement lacks sufficient adaptability according to regional health officials. Healthcare facilities must either follow official procedures or use unofficial procurement methods when facing emergencies or supplier defaults. The system’s inflexibility runs counter to the basic healthcare requirement of immediate access to necessary medications. The lack of an effective monitoring and evaluation (M&E) system creates additional problems for these issues. There exists no standardised reporting system to track non-compliance or delivery delays and suppliers lack consistent accountability measures. The policy’s goals remain unattainable because of administrative weaknesses which prevent decision-makers from making timely adjustments to procurement strategies.

From the qualitative findings, stakeholders showed their understanding of the policy alongside their varied opinions about its execution. The survey results showed that 90% of participants both understood the Framework Agreement and its established provisions. The positive results demonstrate effective policy dissemination together with successful stakeholder education efforts. The majority of pharmacists together with supply chain personnel expressed negative opinions about the policy’s effects. The majority of pharmacists and supply chain personnel described the implementation process as poorly coordinated and top-down while being detached from actual on-the-ground realities. A senior pharmacist expressed concern that the inability to procure outside the Framework Agreement during urgent or emergency situations endangers patients while overburdening health facility operations. Healthcare professionals provided important information which demonstrates a major implementation gap. The policy’s goal to achieve value-for-money procurement and fight corruption remains excellent but frontline staff experience frustration because of execution challenges. Stakeholders reported that the policy focused on fighting procurement corruption while enhancing supply equity, lowering administrative workloads and achieving cost savings through economies of scale. These policy drivers align with global procurement reform initiatives that promote both pooled procurement and long-term contracting approaches for public health sectors. The practical execution of Framework Agreements in the region has not met the expected standards despite their good intentions. The system continues to perform poorly because of multiple systemic issues which include delayed NHIS reimbursements, limited financial independence at lower levels and weak regulatory enforcement. The policy remains ineffective because it failed to adjust to the existing financial problems and emergency stock requirements and regional supply chain limitations. The Framework Agreement system proves useless during crisis situations because regional stores cannot obtain alternative supplies when they run out of stock according to one respondent.

## Conclusion

The study assessed the impact of Framework Agreements on the availability of 65 Essential Medicines at the Upper East Regional Medical Stores. The analysis of both quantitative and qualitative data shows that the Framework Agreement system had good intentions but its implementation in the Upper East Region has resulted in reduced stock availability and longer lead times. The shows that Framework Agreements have negatively affected the availability of essential medicines in the Upper East Region. The FWA policy itself does not contain any fundamental errors but the implementation process has neglected essential operational requirements.

The system’s inflexibility combined with delayed payments and insufficient supplier interaction and non-existent real-time monitoring systems represent major weaknesses that require immediate attention. A procurement system which fails to adapt to health emergencies or local conditions endangers patient outcomes.

### Recommendations

The Ministry of Health and Ghana Health Service should introduce flexibility in procurement rules to permit emergency procurement when contracted suppliers fail or Regional Medical Stores run out of stock. Supplier engagement and coordination should be strengthened through quarterly review meetings and performance dashboards. Payment timeliness should be improved by resolving NHIS payment delays. Monitoring and Evaluation systems should be built with digital real-time tracking and sanctions for defaulters. Certain procurement functions should be decentralised to improve responsiveness. Lastly, there should be regular policy review and stakeholder consultation with inputs from hospitals, regional stores and pharmaceutical suppliers.

## Data Availability

All relevant data are within the manuscript and its Supporting Information files.

## ACKNOWLEDGMENTS

The author expresses sincere gratitude to Dr. Kwaku Gyamfi Oppong for his valuable supervision, guidance and feedback throughout the research process. Appreciation is extended to the staff of the Department of Pharmacy Practice, Kwame Nkrumah University of Science and Technology, for their academic and administrative support. The author also thanks the pharmacists, supply chain officers, and supplier representatives who participated in the study for their time, insights and cooperation.

